# Multimodal Large Language Models vs. Medical Doctors in Degenerative Lumbar Spine Surgery: A Retrospective Decision Concordance Study of 147 Patients

**DOI:** 10.64898/2026.08.04.26359718

**Authors:** Mohammad Hamdan, Anas Al-Bakheet, Imke Fuetterer, Ibrahim Alshaer, Ali Harati

**Author notes:** **Corresponding author:** Mohammad Hamdan, MD. Department of Neurosurgery and Spine Surgery, Fachklinik 360, Ratingen, Germany.

## Abstract

**Objective:** To evaluate decision concordance between commercially available multi-modal large language models (LLMs), resident doctors, and senior-surgeon ground truth for surgical indication and spinal level in degenerative lumbar spine disease.

**Methods:** We retrospectively analyzed 147 consecutive patients. Each case included clinical documentation and MRI presented as two composite PNG images. Two resident doctors and three multimodal LLMs (GPT 5.5, Claude Sonnet 4.6, Gemini 3.1 Pro) independently assessed operative versus conservative management and, if operative, the surgical level. Analyses used Cochran’s Q, McNemar tests with Holm correction, and Bayesian methods.

**Results:** LLMs achieved higher therapy-decision accuracy (66.0%–68.0%; 97–100/147) than residents (54.4%; 80/147) but over-recommended surgery. Conditional level accuracy when surgery was correctly indicated was 71.4% (20/28) for residents versus 33.3%–41.1% for LLMs.

**Conclusion:** Off-the-shelf multimodal LLMs approximate human performance for binary surgical indication but remain inferior for precise level localization. These results establish a practice-relevant baseline of spatial reasoning limitations for tools already used by patients and junior doctors.

## 1 Introduction

Degenerative disorders of the lumbar spine such as lumbar spinal stenosis represent one of the most frequent indications for spinal surgery in the elderly population, while lumbar disc herniation is the leading cause of lumbosacral radiculopathy.[1, 2] Large prospective randomized trials, including the Spine Patient Outcomes Research Trial (SPORT), have demonstrated that appropriately selected patients undergoing surgical treatment achieve superior outcomes compared with nonoperative management for both lumbar disc herniation and lumbar spinal stenosis.[3, 4] However, patient selection remains critical, and treatment decisions continue to require individualized interpretation of heterogeneous clinical and imaging data. As a result, substantial variability in therapeutic recommendations persists among clinicians despite established guidelines and evidence-based treatment pathways.

In recent years large language models (LLMs) have emerged as promising tools for supporting clinical decision-making. These models have demonstrated strong performance in biomedical reasoning, diagnostic tasks, and treatment recommendation across multiple medical domains.[5] In addition, studies assessing the fidelity of medical reasoning have shown that LLMs are increasingly capable of generating structured and clinically coherent decision pathways, although limitations remain regarding hallucinations, calibration of uncertainty, and transparency of reasoning processes.[6] Recent evidence suggests that modern LLMs may approach or even exceed physician-level performance in clinically realistic scenarios, with some studies showing AI performing significantly better than medical doctors.[7] Similar findings have been reported in the field of spinal surgery; however, these prior studies often relied on highly selected and curated cases rather than consecutive real-world patient cohorts.[8, 9]

In spine surgery, early studies have reported promising results for LLMs in clinical decision support, patient counseling, and guideline-based treatment recommendations.[8–12] Furthermore, the recent advent of Multimodal Large Language Models (MLLMs) and Vision-Language Models (VLMs) has extended these capabilities into the visual domain, allowing direct interpretation of radiological imaging. Earlier LLMs were unimodal, making image processing via API difficult. This study is among the first to directly integrate multimodal imaging. However, most prior investigations are based on text-only clinical vignettes, structured questionnaires, or simplified 2D image captioning tasks that struggle to bridge the gap to volumetric 3D clinical data.[13, 14] Consequently, these approaches fail to reflect the complexity of realworld spine care, where therapeutic decisions require simultaneous interpretation of clinical documentation, neurological findings, and multi-planar imaging data.

Our study analyzed 147 real-world cases from a spine surgery center in Germany. Clinical decisions in these cases were made through experienced spine surgeons, thereby representing real clinical expert decision-making. All cases were based on routine clinical documentation, including MRI imaging, reflecting true conditions of everyday clinical practice.

These cases were retrospectively evaluated by two resident doctors rotating in the spine rotation and three large language models (OpenAI’s ChatGPT 5.5, Anthropic’s Claude Sonnet 4.6, and Google’s Gemini 3.1 Pro). We deliberately selected these unspecialized, commercially available models because they are the tools patients and junior doctors are already using; the study therefore aims to quantify a practice-relevant performance baseline, including anatomical and spatial reasoning failures, rather than to validate a custom domain-specific system. Assessments focused on treatment decisions (conservative versus surgical management) and, where surgery was indicated, the selection of the appropriate surgical level. Model outputs were compared with actual clinical decisions and analyzed for concordance with real-world treatment pathways.

## 2 Materials and methods

### 2.1 Study Design and Patient Cohort

This was a retrospective comparative study evaluating the therapeutic decision-making accuracy of Large Language Models (LLMs) compared to medical doctors in lumbar spine pathology. A total of 147 consecutive patients treated at a specialized spine center were included in this study. This consecutive sampling ensures that the cases are representative of routine clinical practice, thus reflecting real-world scenarios. Rather than dividing patients into predefined groups, each patient received their optimal individualized treatment, which served as the reference standard (“ground truth”) for evaluating both the medical doctors and the AI models. The decision of the ground truth was made by a senior spine surgeon. Ultimately, 76 patients received operative treatment and 71 were managed conservatively.

To ensure a homogeneous study population, patients with spinal tumors, acute trauma, infections, and severe deformities were excluded. In addition, no patients with absolute neurosurgical emergencies such as cauda equina syndrome were included. All cases were managed in a non-emergent elective clinical setting under standard clinical conditions.

### 2.2 Data Extraction and Processing

For each patient, comprehensive medical records were reviewed to extract the relevant clinical history and physical examination findings. Concurrently, imaging data was retrieved from the hospital’s Picture Archiving and Communication System (PACS). Specifically, T2-weighted axial and sagittal lumbar magnetic resonance imaging (MRI) series were extracted. To mimic the traditional format of printed radiographic films and constrain the data input for the AI models, the entire MRI series for each patient was consolidated into two distinct composite images (one axial overview and one sagittal overview) and saved as standard PNG files. Examples of these composite images are provided in Figure **??**. This image processing and anonymization was performed using a custom automation script written by the first author. Use of commercially available large language models for case evaluation is documented in this section as required when LLMs contribute to the research workflow.

### 2.3 AI Model Evaluation

Three state-of-the-art Large Language Models (LLMs) were evaluated: GPT 5.5 [15], Gemini 3.1 Pro [16], and Sonnet 4.6 [17]. We intentionally tested unspecialized, commercially available multimodal models rather than domain-fine-tuned or institution-specific systems, because these off-the-shelf tools are precisely those currently accessible to patients and junior doctors in routine practice. The aim was therefore not to optimize a proprietary spine-surgery model, but to establish a critical baseline of real-world performance—and, in particular, of spatial and anatomical reasoning failures—under conditions that mirror present-day clinical use. Each model was queried via its respective direct API endpoint. To facilitate this, the composite MRI images (PNG files) were converted to base64 strings using a custom Python script prior to API transmission. The models were provided with the extracted clinical history, physical examination, and these encoded images. The models were instructed to output a definitive clinical decision: either “Operative” or “Conservative.” If “Operative” was selected, the models were further required to identify the exact surgical level. A strict prompt constraint was applied, explicitly instructing the AI to output the formatted answer without providing further reasoning or explanation (the exact prompt and API parameters are provided as Supplementary material).

### 2.4 Medical Doctor Evaluation

To establish a medical doctor performance baseline, the exact same clinical datasets, comprising the clinical history, physical examination, and the two combined MRI images, were presented to two resident doctors currently rotating in the spine surgery department. The residents were explicitly blinded to the ground truth surgical indication and the original clinical outcomes. They independently assessed the cases and provided their therapeutic recommendations and surgical level planning. In instances where the two resident doctors disagreed on the therapeutic course, a third, highly experienced spine surgeon (independent from the surgeon who established the ground truth) independently reviewed the case to break the tie and establish a medical doctor consensus decision.

### 2.5 Outcome Measures

Primary outcomes were:

- Therapy decision accuracy (surgical vs conservative) compared with the clinical ground truth.
- Level accuracy, defined as correct spinal level identification. Level accuracy was assessed in two predefined ways:
- **Unconditional analysis** across all surgically indicated cases.
- **Conditional analysis** restricted to cases with correctly identified surgical decision.

A fully correct clinical decision required both correct treatment indication and correct level identification.

### 2.6 Statistical Analysis

We compared the diagnostic performance of the LLMs and resident doctors against the expert ground truth using a combination of standard and Bayesian statistical methods.

First, we tested for overall differences in accuracy among all evaluators using Cochran’s Q test. We then performed pairwise comparisons between all evaluators using McNemar’s exact tests. To prevent false positives from making multiple comparisons, we adjusted the resulting p-values using the Holm-Bonferroni method. We also measured how well the different evaluators agreed with each other using Cohen’s Kappa.

In addition to these standard tests, we used an analytical Bayesian approach (employing Beta-Binomial models with neutral Beta(1,1) priors). This allowed us to estimate the true underlying accuracy of each evaluator and calculate 95% Credible Intervals (CrI), giving us a clearer picture of the certainty behind the performance gaps.

All statistical tests evaluated two main outcomes: (1) the accuracy of the overall therapy recommendation (Operative vs. Conservative), and (2) the conditional accuracy of identifying the exact spinal level, evaluated only when surgery was correctly recommended. Additionally, therapy confusion matrices were generated for each evaluator to visually illustrate the distribution of true and false operative and conservative classifications.

## 3 Results

### 3.1 Patient Cohort

A total of 147 patients with degenerative lumbar spine disease were included. Based on expert spine surgeon evaluations, 76 patients had an operative indication and 71 were managed conservatively.

### 3.2 Therapy Recommendation Accuracy

Table 1 summarizes the accuracy of each evaluator in recommending the correct treatment (operative vs. conservative) as well as in identifying the correct spinal level.

**Table 1.** Therapy recommendation and unconditional level identification accuracy with Bayesian posterior estimates (mean and 95% Credible Intervals).

| Model | Therapy Recommendation | | | Unconditional Level Identification ($N = 76$) | | |
| --- | --- | --- | --- | --- | --- | --- |
|  | Correct | Accuracy | Post. Mean [95% CrI] | Correct | Accuracy | Post. Mean [95% CrI] |
| Resident Doctors | 80 | 0.544 | 0.544 [0.463, 0.623] | 21 | 0.276 | 0.280 [0.188, 0.386] |
| GPT 5.5 | 100 | 0.680 | 0.678 [0.601, 0.750] | 23 | 0.303 | 0.307 [0.211, 0.414] |
| Gemini 3.1 Pro | 100 | 0.680 | 0.678 [0.601, 0.750] | 22 | 0.289 | 0.293 [0.200, 0.400] |
| Claude Sonnet 4.6 | 97 | 0.660 | 0.658 [0.580, 0.732] | 16 | 0.211 | 0.215 [0.134, 0.315] |

All three LLMs achieved higher therapy recommendation accuracy than the resident doctors (54.4%). GPT 5.5 and Gemini 3.1 Pro each correctly classified 100 of 147 cases (68.0%), while Claude Sonnet 4.6 was correct in 97 cases (66.0%).

When looking at spinal level identification across all 76 operative cases, accuracies were low for all evaluators. This is largely because evaluators who did not recommend surgery were automatically counted as incorrect for level identification.

### 3.3 Direct Comparisons Between Evaluators

Cochran’s Q test showed a statistically significant difference in therapy accuracy across all evaluators (*Q* = 15.89, *p* = 0.0012). Pairwise testing (Table 2) confirmed that LLMs outperformed resident doctors in therapy recommendation accuracy by approximately 11–13 percentage points (*p* = 0.04 after Holm-Bonferroni correction). No significant differences were found among the three LLMs.

**Table 2.** Pairwise therapy accuracy comparisons: McNemar’s exact test (Holm-adjusted) with Bayesian posterior differences.

| Model A | Model B | McNemar $p$ (Holm) | $P(A > B\vert \text{data})$ | Mean Diff (A-B) | Diff 95% CrI |
| --- | --- | --- | --- | --- | --- |
| Resident Doctors | GPT 5.5 | 0.0396 | 0.0079 | -0.135 | [-0.244, -0.025] |
| Resident Doctors | Gemini 3.1 Pro | 0.0396 | 0.0079 | -0.135 | [-0.244, -0.025] |
| Resident Doctors | Claude Sonnet 4.6 | 0.0396 | 0.0208 | -0.114 | [-0.225, -0.004] |
| GPT 5.5 | Gemini 3.1 Pro | 1.0000 | 0.4996 | -0.000 | [-0.106, 0.105] |
| GPT 5.5 | Claude Sonnet 4.6 | 1.0000 | 0.6425 | 0.020 | [-0.087, 0.126] |
| Gemini 3.1 Pro | Claude Sonnet 4.6 | 1.0000 | 0.6425 | 0.020 | [-0.087, 0.126] |

Agreement between evaluators, measured by Cohen’s Kappa, varied considerably. The two leading LLMs showed substantial agreement with each other (*κ* = 0.724), whereas agreement between the resident doctors and any LLM was only fair to moderate (*κ* ranging from 0.32 to 0.45), reflecting fundamentally different approaches to treatment selection.

### 3.4 Conditional Spinal Level Accuracy: The Key Clinical Finding

The most clinically important finding of this study emerges when examining spinal level accuracy *only among cases where the evaluator correctly recommended surgery*. This conditional analysis reflects the real-world scenario in which a surgeon must not only decide to operate, but must also identify the correct level to avoid wrong-level surgery.

Resident doctors recommended surgery in only 28 of the 76 true operative cases and correctly identified the spinal level in 20 of those 28 cases (71.4%). In contrast, the LLMs recommended surgery in 55–56 cases but identified the correct level in only 15–23 cases (33.3%– 41.1%).

Bayesian analysis (Table 3) strongly supports this finding: the probability that resident doctors have a higher conditional level accuracy than any individual LLM exceeds 99%. While the LLMs recommended surgery more often and therefore achieved higher raw therapy accuracy, they were significantly less reliable in identifying exactly where to operate, a limitation with direct implications for patient safety.

**Table 3.** Conditional spinal level accuracy, evaluated only when the evaluator correctly recommended surgery. Bayesian analysis strongly favors medical doctor precision.

| Model | Surgery Recs (TP) | Level Correct | Accuracy | 95% CrI | $P(\text{Res} > \text{Model})$ |
| --- | --- | --- | --- | --- | --- |
| Resident Doctors | 28 | 20 | 0.714 | [0.528, 0.847] | – |
| GPT 5.5 | 56 | 23 | 0.411 | [0.291, 0.542] | 0.995 |
| Gemini 3.1 Pro | 55 | 21 | 0.382 | [0.265, 0.515] | 0.998 |
| Claude Sonnet 4.6 | 45 | 15 | 0.333 | [0.214, 0.480] | 0.999 |

## 4 Discussion

### 4.1 Principal findings

Our findings reveal a fundamental difference in how large language models and medical doctors approach spine surgical decision-making. LLMs such as GPT 5.5 and Gemini 3.1 Pro outperformed the resident doctors on therapy recommendation accuracy (68% vs 54%), but showed a tendency to over-recommend surgery. When faced with the more demanding task of identifying the exact spinal level, the LLMs fell significantly short. Among cases where the evaluator correctly recommended surgery, resident doctors identified the correct spinal level in 71.4% of cases, compared with 33%–41% for the LLMs. This dissociation between binary surgical indication and reliable spinal-level localization is at the heart of our findings and has direct patient-safety implications.

### 4.2 Self-critique: strengths and weaknesses

An important caveat when interpreting the conditional accuracy of resident doctors is selection bias: residents recommended surgery in only 28 of the 76 true operative cases and were likely selecting the most obvious cases. The LLMs, recommending surgery more broadly, faced more ambiguous cases. Comparing across unequal groups may overestimate the true advantage of medical doctors. Future studies should use severity-matched comparisons.

The conservative behavior of the resident doctors should also be understood in the context of this study’s visual limitations. Both the medical doctors and AI evaluators were limited to two static composite PNG images. This constraint likely contributed to the residents’ reluctance to commit to surgery and also exposes a known limitation of current Vision-Language Models in handling 3D volumetric data and spatial reasoning. It is important to note that the study evaluates both groups under identical constraints.

Further limitations include the single-centre retrospective design, ground truth defined by a senior surgeon rather than multi-reader consensus, use of resident doctors rather than board-certified spine surgeons, absence of external multi-centre validation, and a sample size of 147 patients (while substantial for this study, it may be considered modest for broad generalisation).

### 4.3 Relation to previous work

The tendency of the LLMs to over-recommend surgery deserves careful consideration. LLMs are largely trained on published case reports and clinical vignettes, which tend to feature severe pathology requiring operative intervention. This training bias may lower the models’ threshold for recommending surgery and underscores the need for careful clinical calibration before real-world deployment.

Recent real-world evaluations have moved beyond artificial benchmarks to assess LLM performance using authentic clinical cases. Brodeur et al. demonstrated that modern LLMs can achieve performance comparable to experienced physicians under realistic clinical conditions. Prior spine-focused LLM studies have often relied on vignettes or highly selected cases rather than consecutive multimodal cohorts; our design extends that literature by pairing clinical text with imaging under identical constraints for humans and models.

### 4.4 Implications for neurosurgical practice and further work

This study is one of the first to rigorously compare the clinical decision-making of large language models with that of medical doctors in degenerative lumbar spine surgery using real patient data, including both clinical documentation and MRI imaging. By evaluating exactly those tools, this work establishes a critical baseline for their current limitations—most importantly the dissociation between binary surgical indication and reliable spinal-level localization, a spatial reasoning failure with direct patient-safety implications. Future specialty-tuned or guideline-augmented systems should be benchmarked against this baseline rather than assumed to inherit safe anatomical performance from improved therapy classification alone.

A major limitation of this study is the retrospective design. Future efforts should focus on improving the standardization and quality of clinical documentation. AI models may play a crucial role here, assisting in standardizing clinical notes and capturing a more comprehensive anamnesis.

## 5 Conclusion

Current large language models can process multimodal clinical data and recognize the general need for surgical intervention with reasonable accuracy. However, their tendency to over-recommend surgery and their limited precision in identifying the correct operative level mean they are not yet ready to serve as independent decision-making tools in spine surgery. Future efforts to integrate AI into clinical spine practice should focus on improving anatomical localization accuracy and calibrating surgical thresholds, so that these models can complement rather than replace the clinical judgment of experienced surgeons.

**Figure 1.**
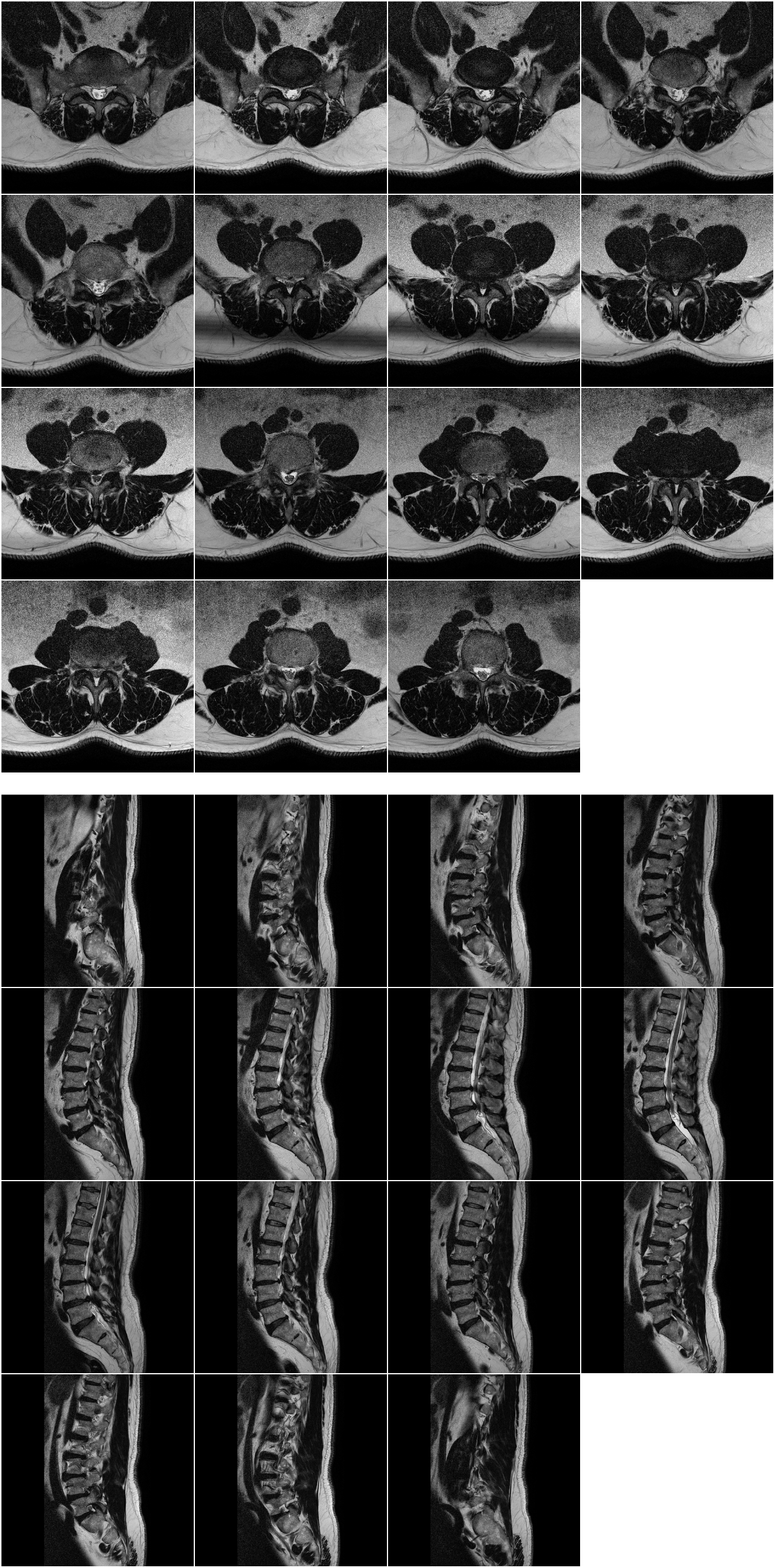
Examples of the composite MRI images (top: axial, bottom: sagittal) presented to both the artificial intelligence models and medical doctors

**Figure 2.**
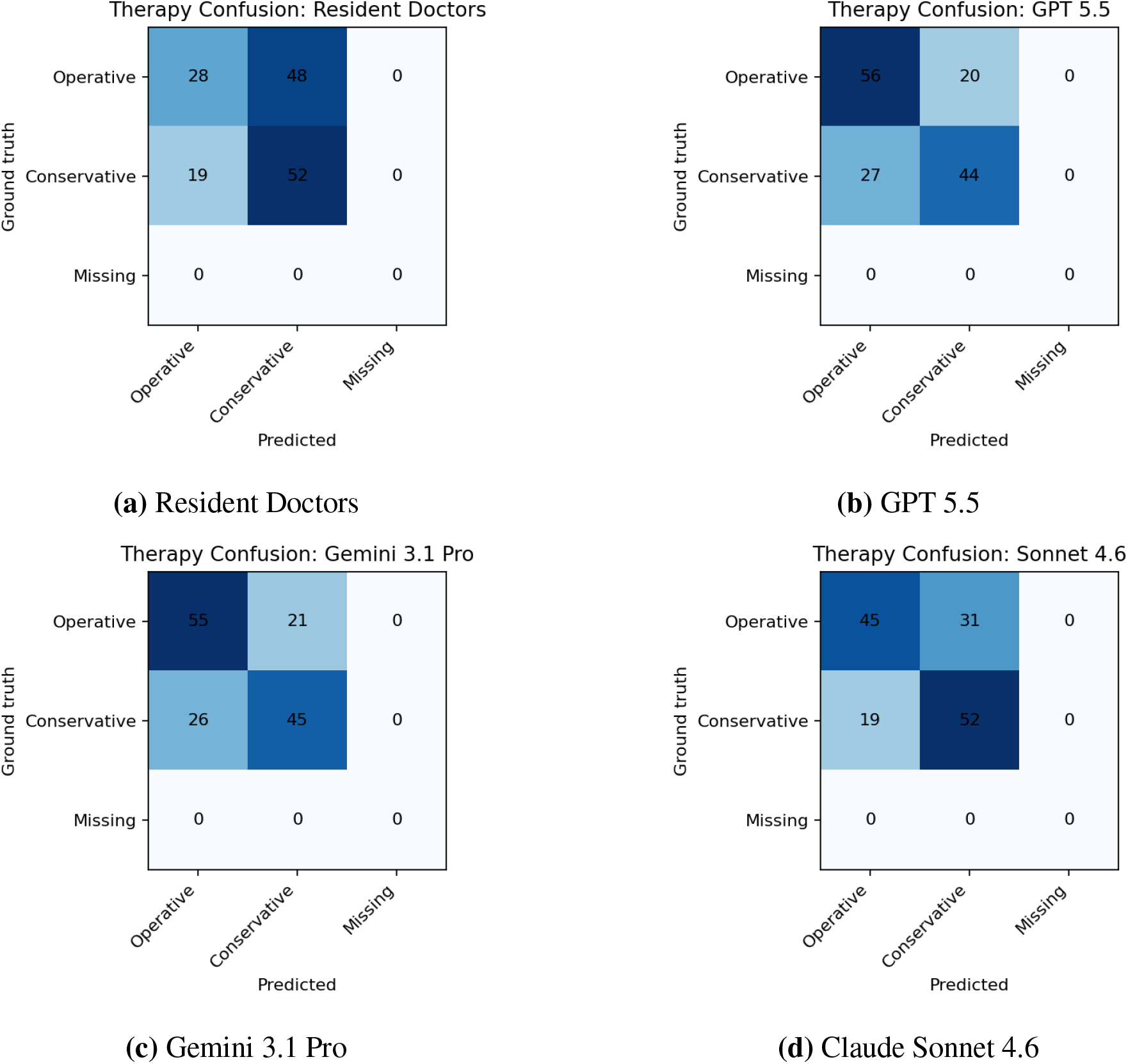
Confusion matrices for therapy decisions. Resident doctors showed a conservative bias (high false-negative rate), while LLMs tended to over-recommend surgery (high false-positive rate).

**Figure 3.**
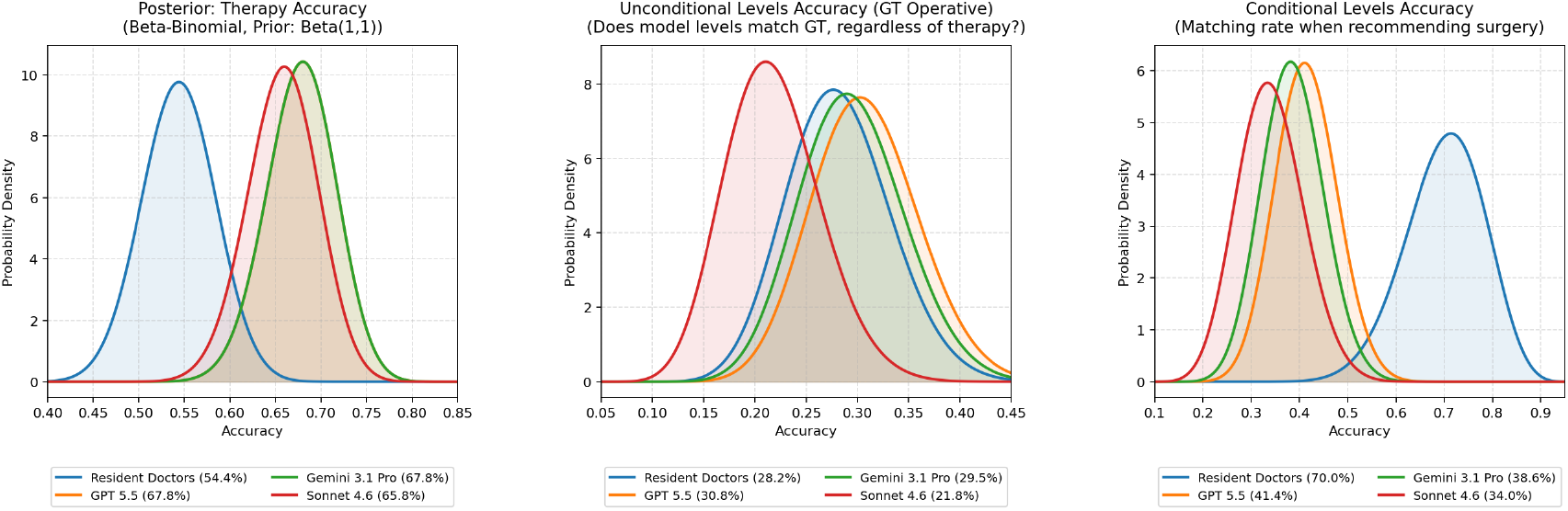
Bayesian posterior probability densities for model accuracies across therapy and level identification metrics.

## Declarations

### Ethics approval

This retrospective study involving human data was conducted in accordance with the ethical standards of the institutional and/or national research committee and with the 1964 Helsinki Declaration and its later amendments or comparable ethical standards (World Medical Association). Ethical approval was waived because the study used only retrospectively collected, anonymized routine-care documentation and imaging; no additional interventions were performed, and patient privacy rights were protected throughout data handling. No ethics approval reference number applies because the study was exempted from formal review on this basis.

### Consent

For this retrospective analysis of fully anonymized routine-care data, formal informed consent to participate was not required under applicable national and institutional rules for non-interventional chart-and-imaging review. The manuscript does not contain case details or images that identify individual participants.

### Data availability

The datasets generated and analyzed during the current study contain sensitive clinical information and are not publicly available due to patient privacy and institutional data-protection regulations. De-identified summary data supporting the findings are available from the corresponding author on reasonable request and with appropriate institutional approval.

### CRediT authorship contribution statement

**Mohammad Hamdan:** Conceptualization, Methodology, Formal analysis, Investigation, Data curation, Writing – original draft, Writing – review & editing. **Anas Al-Bakheet:** Investigation, Data curation, Writing – review & editing. **Imke Fuetterer:** Investigation, Data curation, Writing – review & editing. **Ibrahim Alshaer:** Investigation, Data curation, Writing – review & editing. **Ali Harati:** Conceptualization, Methodology, Formal analysis, Supervision, Writing – review & editing.

### Declaration of generative AI and AI-assisted technologies in the manuscript preparation process

The authors did not use generative AI or AI-assisted technologies in the writing or editing of this manuscript. Large language models were evaluated as study subjects (research tools under test) as described in Materials and Methods; they were not used as authors or as substitute writers of the paper.

